# Action selection in early stages of psychosis: an active inference approach

**DOI:** 10.1101/2022.05.11.22274896

**Authors:** Franziska Knolle, Elisabeth Sterner, Michael Moutoussis, Rick A Adams, Juliet D. Griffin, Joost Haarsma, Hilde Taverne, NSPN Consortium, Ian M. Goodyer, Paul C. Fletcher, Graham K Murray

## Abstract

**Background and Hypothesis:** In order to interact successfully with our environment, we need to build a model, to make sense of noisy and ambiguous inputs. An inaccurate model, as suggested to be the case in psychosis, disturbs optimal action selection. Recent computational models, such as active inference (AI), have emphasized the importance of action selection, treating it as a key part of the inferential process. Based on an AI-framework, we examined prior knowledge and belief precision in an action-based task, given that alterations in these parameters have been linked to the development of psychotic symptoms. We further sought to determine whether task performance and modelling parameters would be suitable for classification of patients and controls.

**Study Design:** 23 at-risk-mental-state individuals, 26 first-episode psychosis patients and 31 controls completed a probabilistic Go/NoGo task in which action choice (Go/ NoGo) was dissociated from outcome valence (gain/ loss). We examined group differences in performance and AI-model parameters, and then performed receiver operating characteristic (ROC) analyses to assess group-classification.

**Study Results:** We found reduced overall performance in patients. AI-modelling revealed that patients showed increased forgetting, reduced confidence in policy selection and less optimal general choice behavior, with poorer action-state associations. Importantly, ROC-analysis revealed fair-to-good classification performances of all groups, when combining modelling parameters and performance measures.

**Conclusion:** Findings show that AI-modelling of this task not only provides further explanation for dysfunctional mechanisms underlying decision making in psychosis, but may also be highly relevant for future research on the development of biomarkers for early identification.

## Introduction

In order to make the most adaptive choices, the brain’s fundamental computational challenge is to integrate sensory data and prior knowledge while taking their uncertainties into account, as both sources (sensory data and prior knowledge) are by themselves inconclusive ^1,2^. This process is formalized as Bayesian inference, in which an initial probabilistic expectation about the state of the environment (the prior) is combined with the probability of the observed sensory data (its likelihood) to compute an updated prediction (the posterior) in which the contributions of prior and likelihood are weighted by their relative precisions ^2,3^. Effective action selection is characterized by maximizing rewards, and minimizing losses through forming accurate beliefs. Thus, in addition to inferring the current state of the environment, the brain is also required to consider possible action outcomes to choose the most probable policy, given the organism’s expectations about its goals and about the likely state of the environment.

In psychosis, optimal action selection seems to be impaired as alterations in value-based action selection ^4,5^ (e.g., reinforcement learning) and in action-outcome (reward/punishment) learning can be observed ^6–12^. These behavioural and cognitive alterations often precede disease onset ^13^; and, therefore, are a robust sign of the pathophysiology of the disorder ^14^. Computational models of decision making allow the investigation of whether and how these impairments contribute to positive and negative symptoms. Reinforcement learning (RL) and active inference (AI) are two frameworks that use different algorithms to approximate how the brain optimises action selection ^1^. Associative RL models (such as model-free RL) make the assumption that fairly simple associative updates (mainly based on reward presentation) are able to accommodate complex task structures and that relatively simple update rules allow powerful performance even when the associations are shaped by higher order contingencies ^2,3^. AI models, on the other hand explicitly posit a more complex model of the structure of the task, and use Bayesian inference not just to infer hidden states of the world, but also to plan and select actions. Here, the primary goal of the agent is to minimize ‘surprise’ (e.g., I want to go outside and I don’t want to be too hot), which the agent achieves by acting to bring about sensory inputs that it expects (e.g., if I wear shorts and a T-shirt, I will be nice and cool), or, in Bayesian terms, maximizing evidence for its assumptions about the environment (i.e. model evidence; it is sunny and people walk around in short sleeves, ergo, I am expecting it to be warm, so shorts and t-shirt will keep me cool) given the context, or state the world is in ^3,15^. In other words, the brain predicts the consequences of an action based on both past experiences and the structure of the task, and will then choose the action expected to produce its most preferred outcomes ^16^. When an individual is unable to use sensory information to correct prior beliefs, or when her prior beliefs are inaccurate and the precision of sensory information is inaccurate, psychotic symptoms, especially hallucinations, may arise ^3,15,17,18^. Not only sensory processing may be conceptualized as an inferential process, but also decision-making during which prior beliefs are the basis of inferring hidden states of the world ^19–21^.

In a recent study, we ^22^ investigated whether reinforcement learning in particular acts as an intermediate phenotype between genetic predispositions and the expressed clinical symptoms. We used an orthogonalised Go/NoGo task ^23^, that was designed to ensure that the reward/punishment outcomes of trials were dissociated from whether a Go or NoGo action was required in early psychosis patients (i.e., at-risk mental state for psychosis individuals (ARMS) and first episode psychosis patients (FEP)), and healthy controls. Using a RL modelling algorithm we found that reward and punishment sensitivity was reduced in both ARMS and FEP compared to controls. The RL model of the Go/NoGo task allows to study computational processes of the brain that mechanistically underlie and lead to specific task performances. AI, while more complex, has some potential advantages over existing RL models of this task ^23–28^: i) it incorporates task structure (as a ‘model-based’ RL agent also would), ii) it treats apparent biases towards certain actions (e.g. ‘no-go’ in negatively valanced states) as prior beliefs rather than fixed action-selection biases, making them easier for agents to overcome, and iii) it can update the confidence with which it chooses actions (its ‘policy precision’) and thus become less random in its choices as it learns more about the task (see ^1^ for further discussion). Indeed, it has been suggested that psychotic symptoms occur as a result of an imbalance between the precision of prior beliefs in relation to sensory evidence ^3,17,29^. Using the same task as Montagnese and co-workers, ^22^, Adams et al. ^1^ showed that an AI model outperformed the RL models in the better-performing individuals.

Cognitive dysfunction, such as dysfunction in decision making, is a core feature of psychosis and has been found to predict poor functional and clinical treatment outcomes ^30^. In at-risk individuals levels of cognitive impairment have been reported to be intermediate compared to healthy controls and schizophrenia ^14^, however, without clear evidence for subsequent decline ^31^. Investigating cognitive impairments in decision making using a modelling approach in at-risk individuals may, therefore, allow the identification of a potential biomarker of risk that improves early identification and intervention ^32,33^.

In this study, we aimed to investigate whether AI parameters of the modelled orthogonalized Go/NoGo task ^23^ differ between ARMS individuals, FEP patients and healthy individuals, and whether they are linked to symptoms. Furthermore, we wished to explore whether task performance and modelling parameters would be suitable for classification of patients and controls.

## Methods

### Participants

The participants have been drawn from the NCAAPS Psychosis dataset which consists of three groups aged 17 to 35, with a total of *N* = 31 Controls, *N* = 23 At-Risk Mental State (ARMS) and *N* = 26 First-Episode Psychosis (FEP). Participants were recruited from the wider population of Cambridgeshire. FEP participants were recruited from the Cambridge First Episode Psychosis service, CAMEO. Classification of ARMS individuals was based on the Comprehensive Assessment for At Risk Mental States (CAARMS, ^34^) as described in Morrison et al. ^35^. All ARMS participants met CAARMS attenuated psychotic symptoms criteria. Controls were recruited through advertisement in Cambridgeshire and through existing University of Cambridge research databases. For a detailed description of selection and classification see Montagnese et al. ^22^.

Table 1 describes participants, including medication status, who were included in all analyses. The study was approved by the Cambridgeshire 3 National Health Service research ethics committee. All subjects gave written informed consent in accordance with the Declaration of Helsinki.

**Table 1.**
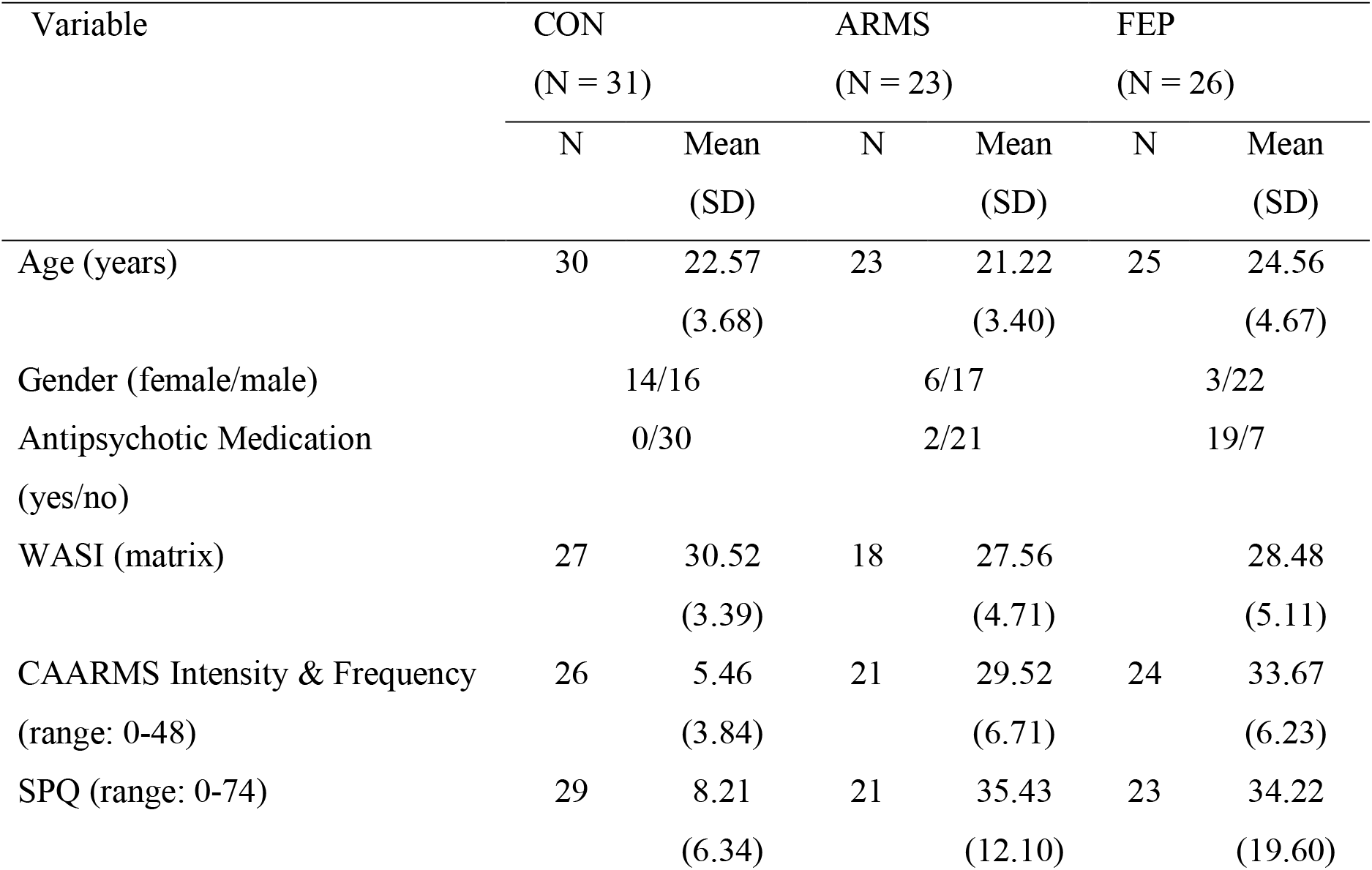
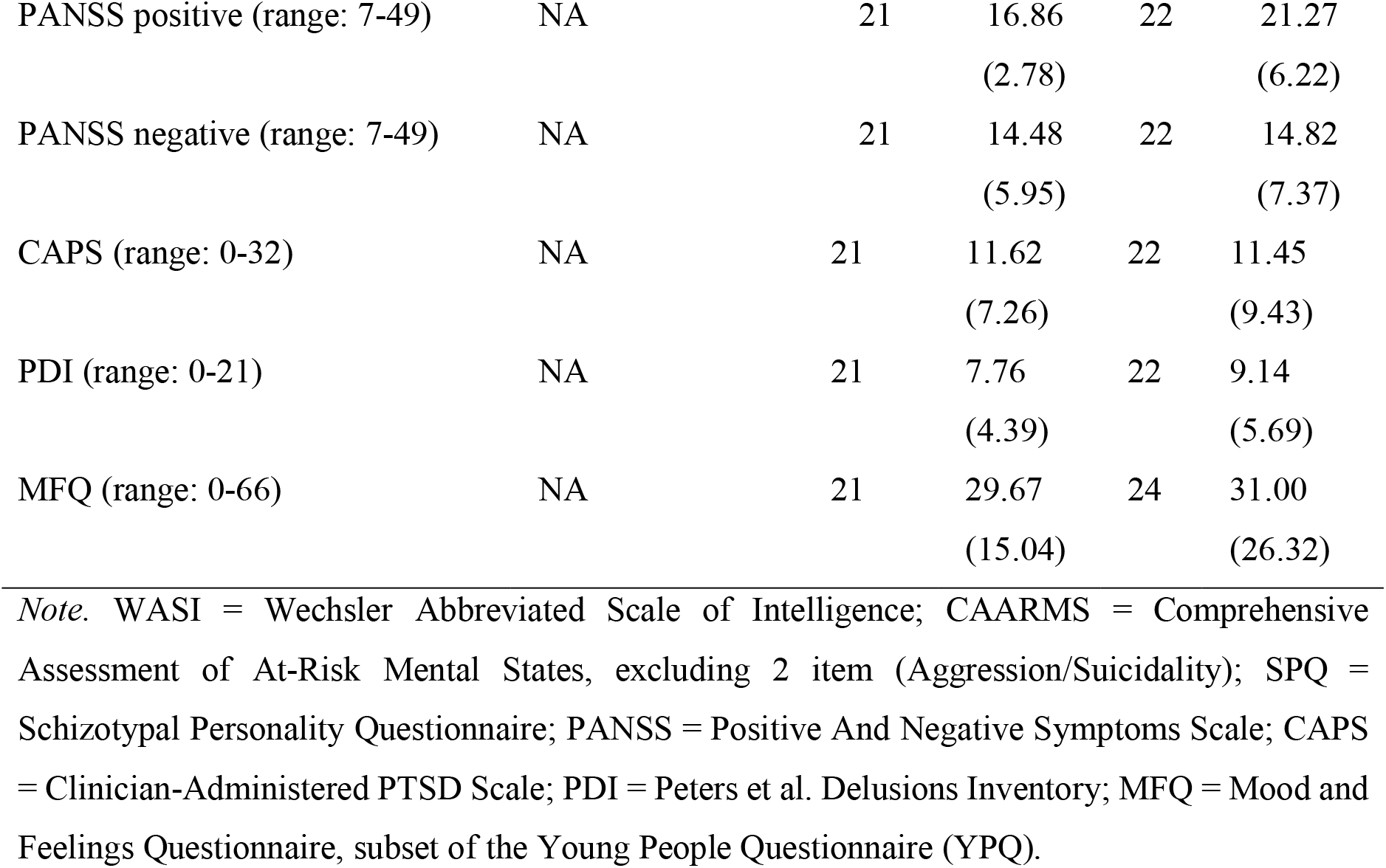
Summary of Demographic Information and Clinical Measures by Group

### Psychological and clinical measures

See Montagnese et al. ^22^ for description of all measures assessed. The relevant measures for the present study are the matrix subscore of the Wechsler Abbreviated Scale of Intelligence (WASI; ^36^), the Comprehensive Assessment of At-Risk Mental State Questionnaire (CAARMS 37); the Schizotypical Personality Questionnaire (SPQ; ^38^), the Positive and Negative Symptoms Scale (PANSS; ^39^), the Clinician-Administered PTSD Scale (CAPS;^40^), the Peters et al. Delusions Inventory (PDI), and the Mood and Feelings Questionnaire (MFQ; ^41^), subset of the Young People Questionnaire (YPQ, ^41^) to measure depressive symptoms.

### Go/NoGo Task

All participants completed an orthogonalized Go/NoGo task, developed by ^23^ which allows the investigation of learning of state-action contingencies. The task is described in detail elsewhere ^22,23^. In short, on each trial one of four different fractal images is presented randomly, and a response - either a Go or a NoGo - has to be performed. The response leads to a probabilistic outcome; possible outcomes are wins (+£0.5), losses (-£0.5), or no-change (+/-£0). Each of the four images represents one condition but participants do not know this. In the two reward conditions, win or no-change are possible outcomes, while in the two punishment conditions, loss or no-change are possible outcomes. Outcomes are assigned probabilistically (80:20). In each pair of reward and punishment conditions, the decisions to be taken in order to maximize the overall win are opposite: ‘Go-to-Win’ and ‘NoGo-to-Win’ for the reward conditions, and ‘NoGo-to-Avoid-Losing’ and ‘Go-to-Avoid-Losing’ for the punishment conditions. The first of each pair is considered a Pavlovian-congruent condition, the second a Pavlovian-incongruent condition.

### Computational modelling – Active inference

In applying computational modelling to psychological tasks, we aim to identify and estimate parameters that relate to the underlying psychological processes engaged in successful task performance and, thereby, to characterise alterations in the processes in clinical groups.

Here we used an active inference modelling approach of the orthogonalized Go/NoGo task as described in ^1^. This approach uses a partially observable Markov Decision Process in order to model the action-dependent state transitions. The agent has to infer the state she is in, considering all actions and outcomes of the past (subject to forgetting), and simultaneously, to infer optimal actions given the current state and taking into consideration her preferences. See Friston et al. ^42,43^ for details. In our model, Pavlovian behaviour is explained on the basis of prior beliefs: *Go* is more likely to be the correct action given a rewarding (Win) context, and No-Go more likely given a punishment (Avoid Losing) context. The model we used thus contained the following parameters: two Pavlovian priors, one in the context of reward (P(a* = go|context = Win), hereafter: Pavlovian win prior), one in the context of punishment (P(a* = no go|context = AL), hereafter: Pavlovian loss prior); an overall prior that the context is one of reward (P(context=Win), hereafter: optimism prior); the precision of the preferences over outcomes, quantifying how strongly rewards are preferred over losses (hereafter: outcome sensitivity); the prior on policy precision, quantifying confidence in choosing (given one’s knowledge and preferences); and finally forgetting, assessing working memory. Importantly, in the AI model, parameters are estimated based on the complete cohort, consisting of the ARMS, FEP and healthy controls to avoid false-positive biases that can occur in some types of hierarchical model-fitting ^44^. This allowed us to investigate the relationship between model parameters and group classification.

### Analysis

Group differences in performance between the four conditions (Go-to-Win, NoGo-to-Win, NoGo-to-Avoid-Losing, Go-to-Avoid-Losing) were assessed using the within-variables valence (positive vs. negative) and action (invigoration vs. inhibition) in a mixed analysis of variance (ANOVA) design with subsequent Tukey post hoc tests.

One-way ANOVAs and Tukey post hoc tests were performed to compare AI parameters between groups. To investigate whether the AI model provides a better fit for individuals who performed above chance in the last 20 trials each condition as done in Adams et al. (2020), we used these trials to assign participants to groups of ‘learners’ (above chance in all conditions) or ‘non-learners’ and then used a two-way ANOVA and Tukey post hoc analyses to compare model fit as assessed by maximum likelihood measures between groups.

Logistic regression was used to analyse the relationship between AI parameters and group membership. We performed receiver operating characteristic (ROC) analyses, and assessed the area under the curve (AUC) to evaluate whether AI parameters contribute to the classification of individuals. AUC thresholds for classification were defined as follows: excellent = 0.90–1, good = 0.80–0.89, fair = 0.70–0.79, poor = 0.60–0.69, or fail = 0.50–0.59 (Safari et al., 2016).

Finally, we used exploratory heatmaps reporting Pearson’s correlation analyses to link AI parameters with clinical measures.

All statistical analyses were conducted in R ^45^. Data was visualized using the ggplot2 package 3.3.5 ^46^. To compute Levene’s test for homogeneity of variance, the car package 3.0-11 ^47^ was used. ANOVA analyses were completed using the afex package 1.0-1 ^48^. Post hoc tests were performed with the lsmeans 2.30-0 package ^49^. Logistic regressions were implemented with the mlogit 1.1-1 package ^50^. ROC and AUC analyses were completed with the pROC 1.18.0 package ^51^. Correlation analyses were performed and visualized with the ggcorrplot package 0.1.3 ^52^.

## Results

### Behavioural performance

Figure 1A depicts the learning rates of each group for each condition ordered along the axes action and valence. In figure 1B, accuracy values for valence and action are presented in boxplots for each group. Individual learning rates at 10 and 30 trials are presented in **Supplementary Figure 1**.

**Figure 1:**
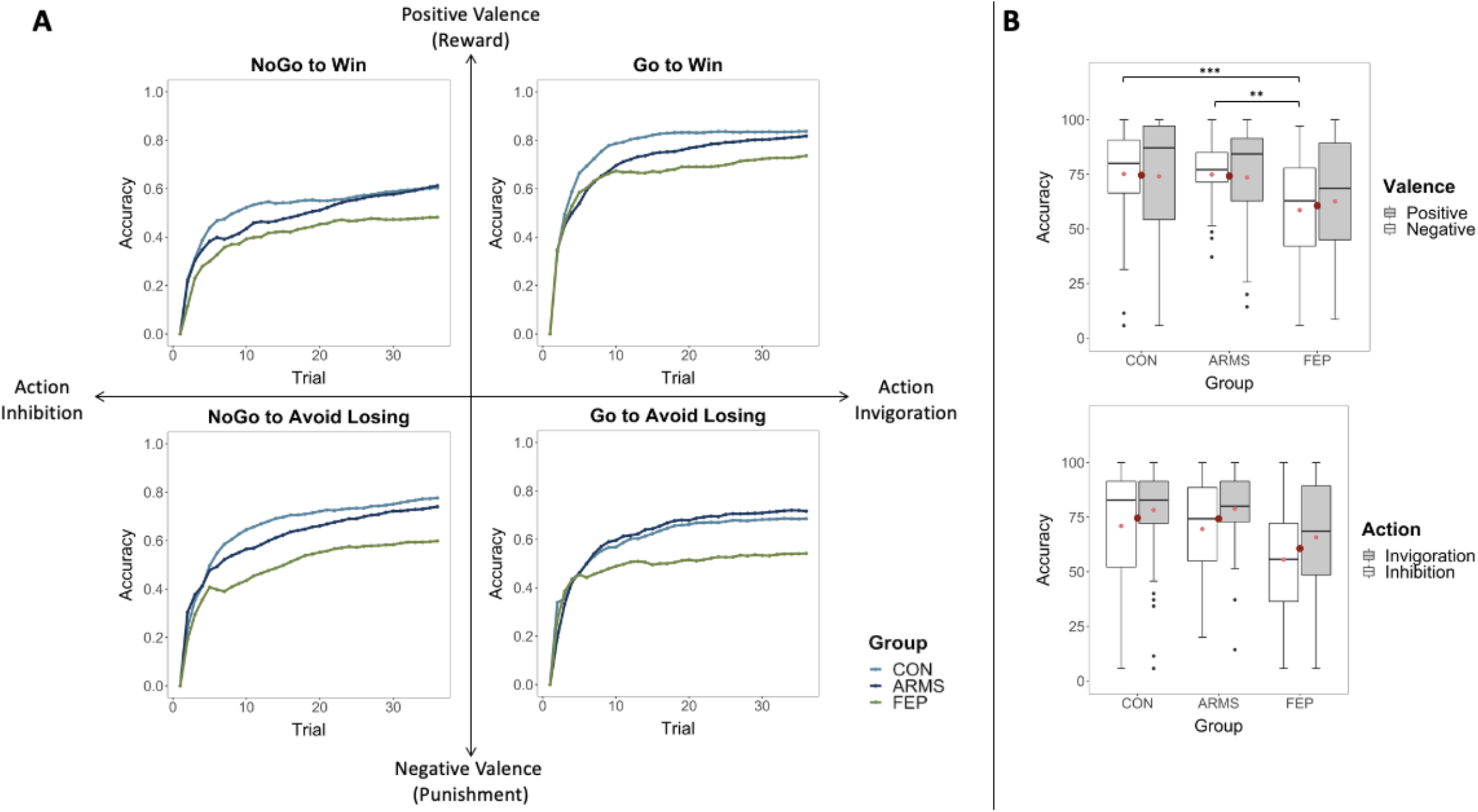
Overview of Task Performance *Note*. A. shows learning rate by group and trial type. Note that FEP participants perform worse (relative to controls) in the Avoid Losing context (lower plots) conditions. B shows behavioural performance. Box plots show the median as horizontal bars, mean as a small red dot, group mean as a large red dot, and interquartile range as whiskers.

Based on this, a 3 × 2 × 2 mixed ANOVA was conducted with group (CON vs. ARMS vs. FEP) as between-group variable and valence (positive vs. negative) and action (invigoration vs. inhibition) as within-group variables. There was a significant main effect for group (*F*(2,77) = 8.02, *p* < .001) and action (*F*(1,77) = 9.30, *p* = .003) but not for valence (*F*(1,77) = 0.15, *p* = .695). While there was no interaction between group and valence (*F*(2,77) = 1.79, *p* = .174) or group and action (*F*(2,77) = 0.09, *p* = .917), there was a significant interaction between action and valence (*F*(1, 77) = 41.65, *p* < .001). This is the Pavlovian effect, as will be discussed below. The group x valence x action interaction was not significant (*F*(2, 77) = 0.42, *p* = 0.660).

Tukey post hoc analysis revealed that Controls (13.98, 95% CI [4.79, 23.16], *p* = .001) and ARMS (13.59, 95% CI [3.70, 23.48], *p* = .004) were significantly more accurate compared to FEP. There was no significant difference in accuracy between Controls and ARMS (0.39, 95% CI [-9.12, 9.89], *p* = 0.995).

Regarding the interaction between action and valence, Tukey post hoc analysis showed that participants were more accurate when performing a Pavlovian congruent response compared to a Pavlovian incongruent response, Go-to-Win compared to Go-to-Avoid-Losing (−15.3, 95% CI [-20.0, -10.7], *p* < .001) and NoGo-to-Avoid-Losing compared to NoGo-to-Win trials (14.3, 95% CI [8.5, 20.1], *p* < .001), respectively.

We also explored the interaction between the within-group variables and group (Figure 1 B). For valence, Tukey post hoc analysis revealed that controls (16.54, 95% CI [5.54, 27.53], *p* < .001) and ARMS (16.28, 95% CI [4.45, 28.11], *p* = .002) were significantly more accurate in punishment trials compared to FEP but not in reward trials. For action, performance did not differ between Go and NoGo trials across Groups.

### Computational Modelling Results

One-way ANOVAs and Tukey post hoc tests were performed to compare AI parameters between groups. We found significant differences for the Pavlovian loss prior, forgetting, as well as for the fit measures Free Energy and Maximum Likelihood (see table 2 and figure 2A for a complete overview of the results).

**Table 2.** Summary of Descriptive Statistics for Active Inference Parameters by Group.

| Parameter | CON |  | ARMS |  | FEP |  | Group<br>Comparison | CON vs. | CON vs. ARMS | ARMS vs. |
| --- | --- | --- | --- | --- | --- | --- | --- | --- | --- | --- |
|  | N | Mean<br>(SD) | N | Mean<br>(SD) | N | Mean<br>(SD) |  | FEP |  | FEP |
| **Optimism prior** | 31 | 0.50<br>(0.05) | 23 | 0.51<br>(0.05) | 26 | 0.52<br>(0.05) | F(2,77) = 0.67,<br>p = .515 |  |  |  |
| **Pavlovian win prior** | 31 | 0.55<br>(0.07) | 23 | 0.55<br>(0.05) | 26 | 0.56<br>(0.07) | F(2,77) = 0.10,<br>p = .901 |  |  |  |
| **Pavlovian loss prior** | 31 | 0.52<br>(0.05) | 23 | 0.50<br>(0.06) | 26 | 0.53<br>(0.05) | F(2,77) = 3.17,<br>p = .048 | -0.01, 95%<br>CI [-0.05,<br>0.02],<br>p = .666). | 0.02, 95% CI<br>[0.01, 0.06],<br>p = .196). | -0.04, 95% CI<br>[-0.07, 0.00],<br>p = .040). |
| **Outcomes sensitivity** | 31 | 5.08<br>(0.10) | 23 | 5.06<br>(0.03) | 26 | 5.04<br>(0.06) | F(2,77) = 0.76,<br>p = .470 |  |  |  |
| **Forgetting** | 31 | 0.41<br>(0.16) | 23 | 0.42<br>(0.21) | 26 | 0.54<br>(0.17) | F(2,77) = 4.95,<br>p = .009 | -0.14, 95%<br>CI [-0.25, -<br>0.02],<br>p = .014). | -0.01, 95% CI [-<br>0.12, 0.11],<br>p = .985). | -0.13, 95% CI<br>[-0.25, 0.01],<br>p = .036). |
| **Prior on Policy**<br>**Precision** | 31 | 1.69<br>(0.65) | 23 | 1.56<br>(0.34) | 26 | 1.35<br>(0.64) | F(2,77) = 2.43,<br>p = .095 | 0.34, 95%<br>CI [-0.03,<br>0.70],<br>p = .078). | 0.12, 95% CI [-<br>0.26, 0.50],<br>p = .723). | 0.21, 95% CI<br>[-0.18, 0.61],<br>p = .399). |
| **Free Energy** | 31 | -62.89<br>(23.04) | 23 | -71.77<br>(14.97) | 24 | -79.53<br>(22.34) | F(2,75) = 4.39,<br>p = .016 | 16.64, 95%<br>CI [3.14,<br>30.14],<br>p = .012). | 8.88, 95% CI [-<br>4.78, 22.55],<br>p = .272). | 7.76, 95% CI<br>[-6.73, 22.24],<br>p = .411). |
| **Maximum Likelihood** | 31 | -60.94<br>(23.16) | 23 | -69.73<br>(14.79) | 26 | -78.58<br>(21.45) | F(2,77) = 5.24,<br>p = .007 | 17.64, 95%<br>CI [4.61,<br>30.68],<br>p = .005). | 8.79, 95% CI [-<br>4.70, 22.29],<br>p = .270). | 8.85, 95% CI<br>[-5.18, 22.89],<br>p = .293). |

**Figure 2:**
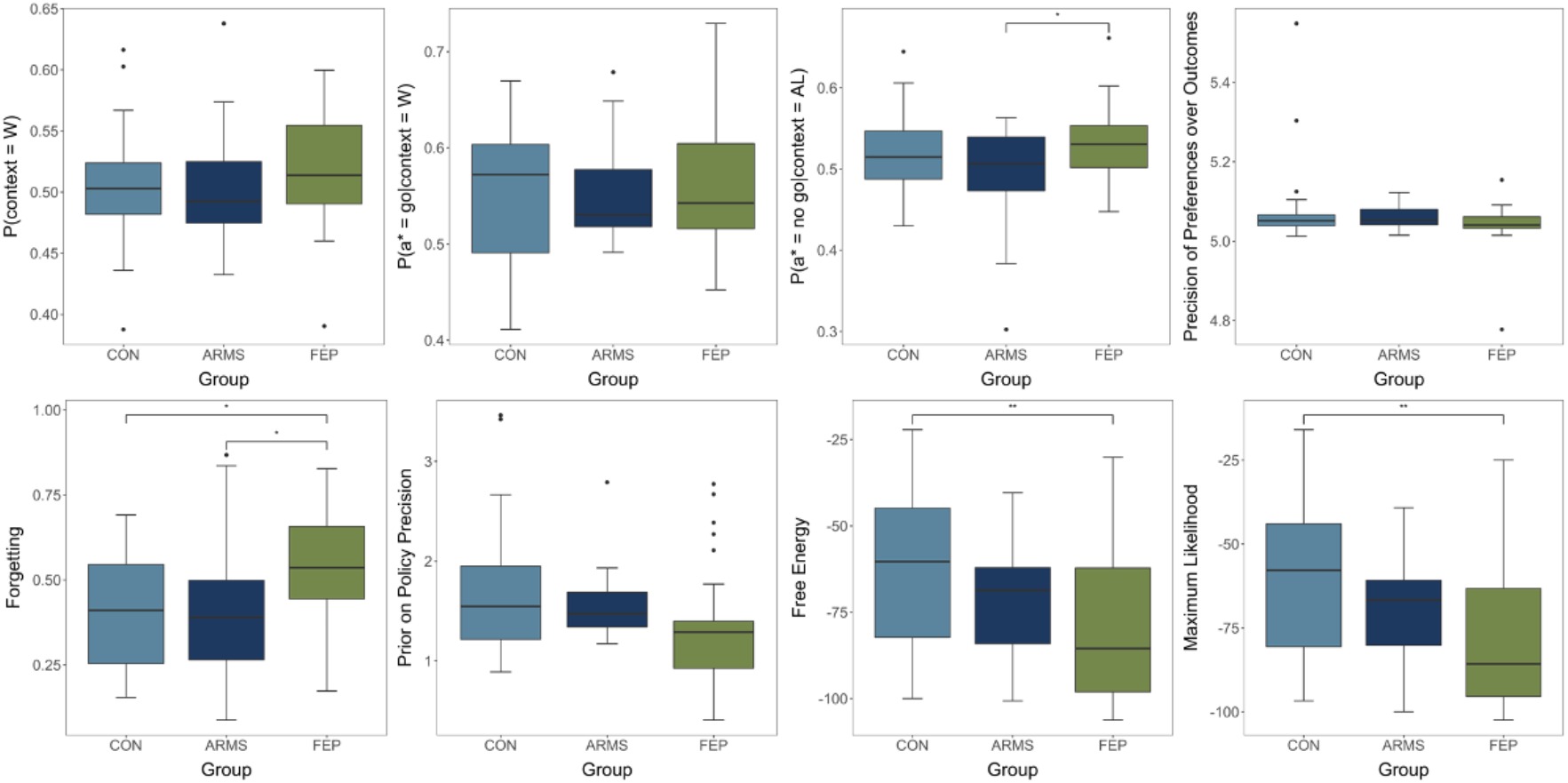
Overview of Group Comparisons of Active Inference Parameters and Model Fit Note. ANOVA analysis of group differences of modelled parameters. Significant results from the Tukey or Games-Howell corrected post hoc analyses are shown (*p < 0.05, **p < 0.01, ***p< 0.001). CON (Healthy Controls); ARMS (At-Risk for Mental Health); FEP (First-Episode Psychosis). Horizontal bars of boxplots mark the median and whiskers indicate the interquartile range. P(context=w): Optimism prior; P(a*=go|context=w): Pavlovian win prior; P(a*=no go|context=AL): Pavlovian loss prior; Precision of Preference over Outcome: Outcome sensitivity.

### Classification based on model parameters

The AUCs from the ROC analyses, representing the overall classification performance based on logistic regression using the AI modelling parameters (Pavlovian win prior, Pavlovian loss prior, optimism prior, outcome sensitivity, forgetting, prior precision, free energy) only, are presented in **Figure 4A**. Classification performances differed depending on group comparison. The controls were differentiated from ARMS with an overall poor performance (AUCs: 0.67, Specificity: 0.65, Sensitivity: 0.61, Accuracy: 0.63). Importantly, the controls were differentiated from FEP with a good performance (AUCs: 0.79, Specificity: 0.83, Sensitivity: 0.68, Accuracy: 0.75), as well as ARMS from FEP (AUCs: 0.82, Specificity: 0.67, Sensitivity: 0.83, Accuracy: 0.74), indicating that AI model parameters contribute significantly to group classification. For regression results see **Supplementary Materials**.

**Figure 4:**
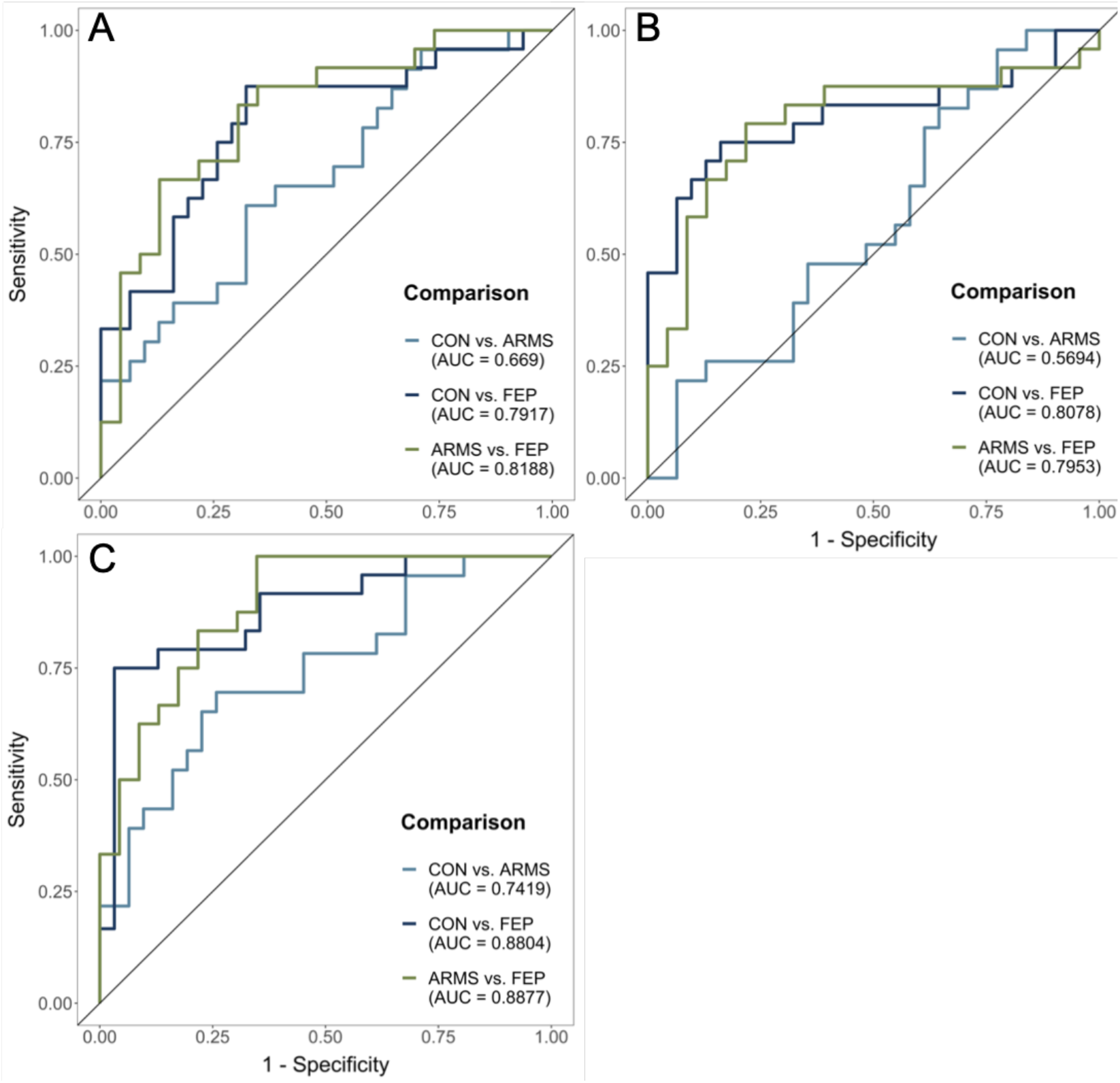
ROC Curves for Group Comparisons Note. ROC curves based on classification models using (A) modelling parameters, (B) performance measures, and (C) a combination of modelling parameters and performance measures. Light blue: CON vs. ARMS; dark blue: CON vs. FEP; green: ARMS vs. FEP. CON: controls; ARMS: individuals at-risk mental state for developing psychosis; FEP: first episode psychosis patients.

### Classification based on performance

The AUCs from the ROC analyses, representing the overall classification performance based on logistic regression using the performance in in the four conditions (Go-to-Win, NoGo-to-Win, NoGo-to-Avoid-Losing, Go-to-Avoid-Losing) only, are presented in **Figure 4B**. Classification performances differed depending on group comparison. The classification of controls and ARMS failed using the performance (AUCs: 0.57, Specificity: 0.48, Sensitivity: 0.58, Accuracy: 0.54). Importantly, the controls were differentiated from FEP with a good performance (AUCs: 0.81, Specificity: 0.75, Sensitivity: 0.71, Accuracy: 0.73), as well as ARMS from FEP (AUCs: 0.80, Specificity: 0.79, Sensitivity: 0.70, Accuracy: 0.74). For regression results see **Supplementary Materials**.

### Classification based on model parameters and performance

The AUCs from the ROC analyses, representing the overall classification performance based on logistic regression using the performance in in the four conditions (Go-to-Win, NoGo-to-Win, NoGo-to-Avoid-Losing, Go-to-Avoid-Losing) and the AI modelling parameters (Pavlovian win prior, Pavlovian loss prior, optimism prior, outcome sensitivity, forgetting, prior precision, free energy), are presented in **Figure 4C**. Classification performances differed depending on group comparison. Importantly, the controls were differentiated from FEP with a good performance (AUCs: 0.88, Specificity: 0.79, Sensitivity: 0.81, Accuracy: 0.80), as well as ARMS from FEP (AUCs: 0.89, Specificity: 0.75, Sensitivity: 0.83, Accuracy: 0.79). In contrast to the performance-only classification’s failure to differentiate these two groups, the controls were differentiated from ARMS with an overall fair performance (AUCs: 0.74, Specificity: 0.70, Sensitivity: 0.65, Accuracy: 0.67). For regression results see **Supplementary Materials**.

### Relationship between AI Parameters and Clinical Parameters

In an exploratory correlation analysis between modelling parameters and clinical scores **(Supplementary Figure 2)**, we found a significant negative relationship between CAARMS and policy precision (*r* = -.28, *p* = .016), Free Energy (*r* = -.38, *p* = .001), and maximum likelihood (*r* = -.40, p < .001) across all participants (**Supplementary Figure 2D**). These correlations are driven by the control group (**Supplementary Figure 2A)**.

## Discussion

The present study investigated whether Bayesian measures of decision making underlying action selection differ in ARMS and FEP compared to healthy controls, and whether they are associated with symptoms. We applied an AI model ^1^ to an orthogonalized Go/NoGo task ^23^ in ARMS individuals, FEP patients, and in healthy controls ^22^. Our results revealed significantly worse performance in the punishment condition in FEP compared to ARMS and controls. Furthermore, FEP had an increased ‘forgetting’ parameter relative to the other groups. Investigating the Bayesian model measures, we found that FEP compared to controls showed significantly lower levels in free energy and maximum likelihood, indicating that patients were less Bayesian-optimal in their inferences than the other groups.

Interestingly, we found higher reliance on the prior in the context of avoiding losing, when no-go is the best action, in FEP compared to ARMS, as well as a trend towards lower prior precision of policies in FEP compared to controls. This is consistent with current theories of an imbalance of prior precision compared to sensory likelihood underlying the psychopathology of symptoms in psychosis. It may also reconcile the findings of Moutoussis ^53^ with those of Ermakova 11 in that inconsistent choosing that might be associated with lower policy precision was found in the former study, which had more longer-term psychosis patients than the latter, which focused on ARMS and early psychosis. However, our data shows that there are eight individuals, five of which in the FEP group, with policy precision above group specific standard deviation, suggesting greater variability with respect to this measure, especially in the FEP group.

Importantly, this task disentangles the type of action (Go vs. NoGo) from the valence of the action outcome (reward vs. punishment). Consistent with prior research using this task ^23,25^, participants generally performed better in the Pavlovian congruent conditions (Go-to-Win, NoGo-to-Avoid-Losing) compared to the incongruent ones (Go-to-Avoid-Losing, NoGo-to-Win). Previous research discussed impairments in action selection to be an intermediate phenotype between neurobiological /genetic substrates and expressed clinical symptoms, which is why these alterations would be expected to be also present in individuals with increased clinical risk of developing psychosis ^54,55^; however findings are inconsistent ^13^. Our study does not confirm this latter hypothesis. In our sample, we identified poorer performance in FEP compared to controls but also compared to ARMS, mainly in the punishment condition. In a recent study using a probabilistic learning task in early and persistent psychosis, Suetani et al. ^56^ report early psychosis patients are less likely to adapt their behavior after a loss compared to controls, which is indicative of deficits in punishment learning and is similar to our findings.

Using the AI model parameters, we identified differences between FEP and controls as well as ARMS. Interestingly, we found that FEP individuals are less likely to achieve the Bayes-optimal outcome selection, indicated by lower Free Energy and maximum log likelihood. Importantly, we found reduced precision of prior policy in FEP compared to controls and ARMS, and an increase in the (Pavlovian) prior belief that NoGo is the best action in the context of avoiding losing. It has been suggested that psychosis is represented by an imbalance of the precision of the prior relative to the precision of sensory information ^3,15,57–59^, this interaction may be dependent on the hierarchical level, with increased precision of the prior at higher hierarchical levels ^60–62^ and decreased precision at lower hierarchical levels ^29^. The results in our study showed lower prior precision and higher forgetting in FEP, indicating that individuals show deficits in identifying and possibly maintaining the associations between cue and outcome, but especially their probabilities, which may be linked to altered beliefs about environmental volatilities ^60,63,64^.

Adams et al. (2020) reported that prior precision is negatively correlated to D2/3 receptor availability in the limbic striatum, linking greater D2/3 receptor availability to lower precision. Findings of D2/3 receptor availability in psychosis, mainly in the striatum and the thalamus, have been linked to medication status, indicating that antipsychotic naïve individuals have the same D2/3 receptor availability as controls ^65,66^. Some studies, however, also indicate an increased availability before or without treatment (Veselinowvić et al., 2018). Adams et al. (2020) reported that lower D2/3 receptor availability can be cognitively advantageous, as higher tonic dopamine activity may be associated with higher prior precision. We speculate that FEP patients with lower prior precision may have more disordered dopaminergic transmission: this will need further investigation.

In this study, we applied the AI model to the whole sample to reduce the likelihood of false-positive findings ^44,68^. Although this procedure may be slightly less sensitive to group specific differences, by fitting the posterior distribution over performances of all individuals, it allows conservative use of the parameter output for classification analyses. Computational modelling of neurocognitive tasks sheds light on the neuropsychology processes underlying the behavior which may be linked to the psychopathology of the disorder ^2,58^, using model parameters in classification algorithms combines two approaches necessary to improve strategies of identifying individuals early and precisely ^69,70^.

The detection of potential biomarkers for early identification of individuals at risk is one of the goals of today’s research effort. Cognitive impairments are particularly interesting due to the prodromal onset and the impact on functional outcome ^14,30,71–73^. Using ROC analysis, we were able to show that the specific expression of modelling parameters (Figure 4A) as well as the individual performances measures (Figure 4B), allowed a good classification of FEP patients, with significant differentiation from controls and ARMS, indicating that differences in the neuropsychological processes underlying the performance in the Go/NoGo task are relevant for the psychopathology of psychosis ^9,13,19^. Importantly, however, when combining modelling parameters and performance measures in the ROC analysis, the identification of all group associations improved by at least 8%. This improvement was especially important for the distinction of controls and ARMS, which now provided a fair classification performance. This finding is highly relevant for future research on the development of biomarkers for early identification and should be validated in larger testing samples.

In conclusion, this study shows that deficits in probabilistic decision making in an orthogonalized Go/NoGo task in FEP patients are linked to increased forgetting, reduced prior precision and less optimal general choice behavior, with poorer punishment learning. Furthermore, we found that optimal choice behavior correlated with symptom strength, indicating less optimal behavior with increased symptoms across ARMS and FEP. Reduced prior precision in FEP may potentially linked to alterations in tonic striatal dopaminergic activity, which is associated with D2/3 receptor availability. Our study hence replicates previous findings and provides further mechanistic insights about how altered cognitive parameters may lead to dysfunctional decision making in psychosis.

## Data Availability

Data is available upon reasonable request to the authors.

## Acknowledgments

We would like to thank all participants for their time and engagement.

## References

1. Adams, R. A. et al. Variability in Action Selection Relates to Striatal Dopamine 2/3 Receptor Availability in Humans: A PET Neuroimaging Study Using Reinforcement Learning and Active Inference Models. Cereb. Cortex 30, 3573–3589 (2020).

2. Adams, R. A., Huys, Q. J. M. & Roiser, J. P. Computational Psychiatry: towards a mathematically informed understanding of mental illness. J. Neurol. Neurosurg. &amp;amp; Psychiatry 87, 53LP–63 (2016).

3. Sterzer, P. et al. The Predictive Coding Account of Psychosis. Biological Psychiatry (2018) doi:10.1016/j.biopsych.2018.05.015.

4. Waltz, J. A., Wilson, R. C., Albrecht, M. A., Frank, M. J. & Gold, J. M. Differential Effects of Psychotic Illness on Directed and Random Exploration. Comput. psychiatry (Cambridge, Mass.) 4, 18–39 (2020).

5. Gold, J. M. et al. Negative symptoms of schizophrenia are associated with abnormal effort-cost computations. Biol. Psychiatry 74, 130–136 (2013).

6. Deserno, L., Boehme, R., Heinz, A. & Schlagenhauf, F. Reinforcement learning and dopamine in schizophrenia: Dimensions of symptoms or specific features of a disease group? Frontiers in Psychiatry vol. 4 (2013).

7. Gold, J. M., Waltz, J. A., Prentice, K. J., Morris, S. E. & Heerey, E. A. Reward processing in schizophrenia: a deficit in the representation of value. Schizophr Bull 34, 835–847 (2008).

8. Morris, R. W., Cyrzon, C., Green, M. J., Le Pelley, M. E. & Balleine, B. W. Impairments in action–outcome learning in schizophrenia. Transl. Psychiatry 8, 54 (2018).

9. Kesby, J. P., Murray, G. K. & Knolle, F. Neural circuitry of salience and reward processing in psychosis. Biol. Psychiatry Glob. Open Sci. (2021).

10. Ermakova, A. O. et al. Abnormal reward prediction-error signalling in antipsychotic naive individuals with first-episode psychosis or clinical risk for psychosis. Neuropsychopharmacology 1 (2018).

11. Ermakova, A. O. et al. Cost evaluation during decision-making in patients at early stages of psychosis. Comput. Psychiatry 3, 18–39 (2019).

12. Maia, T. V & Frank, M. J. An Integrative Perspective on the Role of Dopamine in Schizophrenia. Biol. Psychiatry 81, 52–66 (2017).

13. Strauss, G. P. et al. Reinforcement learning abnormalities in the attenuated psychosis syndrome and first episode psychosis. Eur. Neuropsychopharmacol. 47, 11–19 (2021).

14. Fusar-Poli, P. et al. Cognitive Functioning in Prodromal Psychosis: A Meta-analysis. Arch. Gen. Psychiatry 69, 562–571 (2012).

15. Adams, R., Stephan, K., Brown, H., Frith, C. & Friston, K. The Computational Anatomy of Psychosis. Frontiers in Psychiatry vol. 4 47 (2013).

16. Smith, R., Friston, K. J. & Whyte, C. J. A step-by-step tutorial on active inference and its application to empirical data. J. Math. Psychol. 107, 102632 (2022).

17. Benrimoh, D., Parr, T., Adams, R. A. & Friston, K. Hallucinations both in and out of context: an active inference account. PLoS One 14, e0212379 (2019).

18. Benrimoh, D., Parr, T., Vincent, P., Adams, R. A. & Friston, K. Active inference and auditory hallucinations. Comput. Psychiatry (Cambridge, Mass.) 2, 183 (2018).

19. Sterzer, P., Voss, M., Schlagenhauf, F. & Heinz, A. Decision-making in schizophrenia: A predictive-coding perspective. Neuroimage 190, 133–143 (2019).

20. Friston, K. The free-energy principle: a unified brain theory? Nat. Rev. Neurosci. 11, 127–138 (2010).

21. Botvinick, M. & Toussaint, M. Planning as inference. Trends Cogn. Sci. 16, 485–488 (2012).

22. Montagnese, M. et al. Reinforcement learning as an intermediate phenotype in psychosis? Deficits sensitive to illness stage but not associated with polygenic risk of schizophrenia in the general population. Schizophr. Res. (2020).

23. Guitart-Masip, M. et al. Go and no-go learning in reward and punishment: interactions between affect and effect. Neuroimage 62, 154–166 (2012).

24. de Boer, J. N. et al. Auditory hallucinations, top-down processing and language perception: a general population study. Psychol. Med. 49, 2772–2780 (2019).

25. Huys, Q. J. M. et al. Disentangling the roles of approach, activation and valence in instrumental and pavlovian responding. PLoS Comput. Biol. 7, e1002028 (2011).

26. Swart, J. C. et al. Catecholaminergic challenge uncovers distinct Pavlovian and instrumental mechanisms of motivated (in)action. Elife 6, e22169 (2017).

27. Chowdhury, R., Guitart-Masip, M., Lambert, C., Dolan, R. J. & Düzel, E. Structural integrity of the substantia nigra and subthalamic nucleus predicts flexibility of instrumental learning in older-age individuals. Neurobiol. Aging 34, 2261–2270 (2013).

28. Cavanagh, J. F., Eisenberg, I., Guitart-Masip, M., Huys, Q. & Frank, M. J. Frontal theta overrides pavlovian learning biases. J. Neurosci. 33, 8541–8548 (2013).

29. Weilnhammer, V. et al. Psychotic experiences in schizophrenia and sensitivity to sensory evidence. Schizophr. Bull. 46, 927–936 (2020).

30. Allott, K., Liu, P., Proffitt, T.-M. & Killackey, E. Cognition at illness onset as a predictor of later functional outcome in early psychosis: systematic review and methodological critique. Schizophr. Res. 125, 221–235 (2011).

31. Bora, E. & Murray, R. M. Meta-analysis of cognitive deficits in ultra-high risk to psychosis and first-episode psychosis: do the cognitive deficits progress over, or after, the onset of psychosis? Schizophr. Bull. 40, 744–755 (2014).

32. Fusar-Poli, P. Predicting Psychosis. Arch. Gen. Psychiatry 69, 220 (2012).

33. Velthorst, E. et al. Potentially important periods of change in the development of social and role functioning in youth at clinical high risk for psychosis. Dev. Psychopathol. 30, 39–47 (2018).

34. Yung, A. R. et al. Mapping the onset of psychosis: The comprehensive assessment of at risk mental states. Schizophr. Res. 39, 964–971 (2005).

35. Morrison, A. P. et al. Early detection and intervention evaluation for people at risk of psychosis: multisite randomised controlled trial. Br. Med. J. 344, e2233 (2012).

36. Wechsler, D. Wechsler abbreviated scale of intelligence--. (1999).

37. Yung, A. R. et al. Mapping the onset of psychosis: The Comprehensive Assessment of At-Risk Mental States. Aust. N. Z. J. Psychiatry 39, 964–971 (2005).

38. Raine, A. The SPQ: A Scale for the Assessment of Schizotypal Personality Based on DSM-III-R Criteria. Schizophr. Bull. 17, 555–564 (1991).

39. Kay, S. R., Fiszbein, A. & Opler, L. A. The positive and negative syndrome scale (PANSS) for schizophrenia. Schizophr. Bull. 13, 261–76 (1987).

40. Bell, V., Halligan, P. W. & Ellis, H. D. The Cardiff Anomalous Perceptions Scale (CAPS): A new validated measure of anomalous perceptual experience. Schizophr. Bull. 32, 366–377 (2006).

41. Costello, E. J. & Angold, A. Scales to assess child and adolescent depression: checklists, screens, and nets. J. Am. Acad. Child Adolesc. Psychiatry 27, 726–737 (1988).

42. Friston, K. Active inference and free energy. Behav. Brain Sci. 36, 212 (2013).

43. Friston, K. et al. The anatomy of choice: active inference and agency. Front. Hum. Neurosci. 7, 598 (2013).

44. Moutoussis, M., Hopkins, A. K. & Dolan, R. J. Hypotheses About the Relationship of Cognition With Psychopathology Should be Tested by Embedding Them Into Empirical Priors. Front. Psychol. 9, (2018).

45. Rstudio, T. RStudio: Integrated Development for R. Rstudio Team, PBC, Boston, MA URL http://www.rstudio.com/ (2020) xdoi:10.1145/3132847.3132886.

46. Wickham, H. ggplot2: elegant graphics for data analysis. (springer, 2016).

47. Fox, J. & Weisberg, S. An R companion to applied regression. (Sage publications, 2018).

48. Singmann, H., Bolker, B., Westfall, J., Aust, F. & Ben-Shachar, M. S. afex: Analysis of factorial experiments. R Packag. version 0. 13–145 (2015).

49. Lenth, R. V. Least-squares means: the R package lsmeans. J. Stat. Softw. 69, 1–33 (2016).

50. Croissant, Y. Estimation of Random Utility Models in R: The mlogit Package. J. Stat. Softw. 95, 1–41 (2020).

51. Robin, X. et al. pROC: an open-source package for R and S+ to analyze and compare ROC curves. BMC Bioinformatics 12, 77 (2011).

52. Kassambara, A. & Kassambara, M. A. Package ‘ggcorrplot’. R Packag. version 0.1 3, (2019).

53. Moutoussis, M., Bentall, R. P., El-Deredy, W. & Dayan, P. Bayesian modelling of Jumping-to-Conclusions bias in delusional patients. Cogn. Neuropsychiatry 16, 422–447 (2011).

54. Millman, Z. B. et al. Evidence of reward system dysfunction in youth at clinical high-risk for psychosis from two event-related fMRI paradigms. Schizophr. Res. 226, 111–119 (2020).

55. Waltz, J. et al. Reinforcement learning performance and risk for psychosis in youth. J. Nerv. Ment. Dis. 203, 919 (2015).

56. Suetani, S. et al. Impairments in goal-directed action and reversal learning in a proportion of individuals with psychosis: evidence for differential phenotypes in early and persistent psychosis. medRxiv (2021).

57. Corlett, P. R., Frith, C. D. & Fletcher, P. C. From drugs to deprivation: A Bayesian framework for understanding models of psychosis. Psychopharmacology vol. 206 515–530 (2009).

58. Heinz, A. et al. Towards a Unifying Cognitive, Neurophysiological, and Computational Neuroscience Account of Schizophrenia. Schizophr. Bull. (2019) doi:10.1093/schbul/sby154.

59. Haarsma, J., Kok, P. & Browning, M. The promise of layer-specific neuroimaging for testing predictive coding theories of psychosis. Schizophr. Res. (2020) doi:https://doi.org/10.1016/j.schres.2020.10.009.

60. Haarsma, J. et al. Precision weighting of cortical unsigned prediction error signals benefits learning, is mediated by dopamine, and is impaired in psychosis. Mol. Psychiatry (2020) doi:10.1038/s41380-020-0803-8.

61. Teufel, C. et al. Shift toward prior knowledge confers a perceptual advantage in early psychosis and psychosis-prone healthy individuals. Proc. Natl. Acad. Sci. U. S. A. 112, 13401–13406 (2015).

62. Haarsma, J. et al. Influence of prior beliefs on perception in early psychosis: Effects of illness stage and hierarchical level of belief. J. Abnorm. Psychol. (2020) doi:10.1037/abn0000494.

63. Culbreth, A. J., Westbrook, A., Xu, Z., Barch, D. M. & Waltz, J. A. Intact Ventral Striatal Prediction Error Signaling in Medicated Schizophrenia Patients. Biol. Psychiatry Cogn. Neurosci. Neuroimaging (2016) doi:10.1016/j.bpsc.2016.07.007.

64. Schlagenhauf, F. et al. Striatal dysfunction during reversal learning in unmedicated schizophrenia patients. Neuroimage 89, 171–180 (2014).

65. Chen, K. C. et al. Striatal dopamine D2/3 receptors in medication-naïve schizophrenia: an [123I] IBZM SPECT study. Psychol. Med. 1–9 (2021) doi:DOI: 10.1017/S0033291720005413.

66. Rajji, T. K. et al. Cognition and Dopamine D2 Receptor Availability in the Striatum in Older Patients with Schizophrenia. Am. J. Geriatr. Psychiatry 25, 1–10 (2017).

67. Veselinović, T. et al. The role of striatal dopamine D2/3 receptors in cognitive performance in drug-free patients with schizophrenia. Psychopharmacology (Berl). 235, 2221–2232 (2018).

68. Moutoussis, M. et al. Neural activity and fundamental learning, motivated by monetary loss and reward, are intact in mild to moderate major depressive disorder. PLoS One 13, e0201451 (2018).

69. Wiecki, T. V, Poland, J. & Frank, M. J. Model-based cognitive neuroscience approaches to computational psychiatry: clustering and classification. Clin. Psychol. Sci. 3, 378–399 (2015).

70. Huys, Q. J. M., Maia, T. V & Frank, M. J. Computational psychiatry as a bridge from neuroscience to clinical applications. Nat. Neurosci. 19, 404–413 (2016).

71. Fett, A. J. et al. Long-term Changes in Cognitive Functioning in Individuals With Psychotic Disorders: Findings From the Suffolk County Mental Health Project. JAMA Psychiatry 77, 387–396 (2020).

72. Savla, G. N., Vella, L., Armstrong, C. C., Penn, D. L. & Twamley, E. W. Deficits in domains of social cognition in schizophrenia: a meta-analysis of the empirical evidence. Schizophr. Bull. 39, 979–992 (2013).

73. Guo, J. Y. et al. Predicting psychosis risk using a specific measure of cognitive control: a - 12-month longitudinal study. Psychol. Med. 50, 2230–2239 (2020).

