## Supplementary Materials for "Action selection in early stages of psychosis: an active inference approach"

Running title: Action selection in early psychosis

Knolle et al.

1. **Individual learning rates**

**Supplementary Figure 1:**

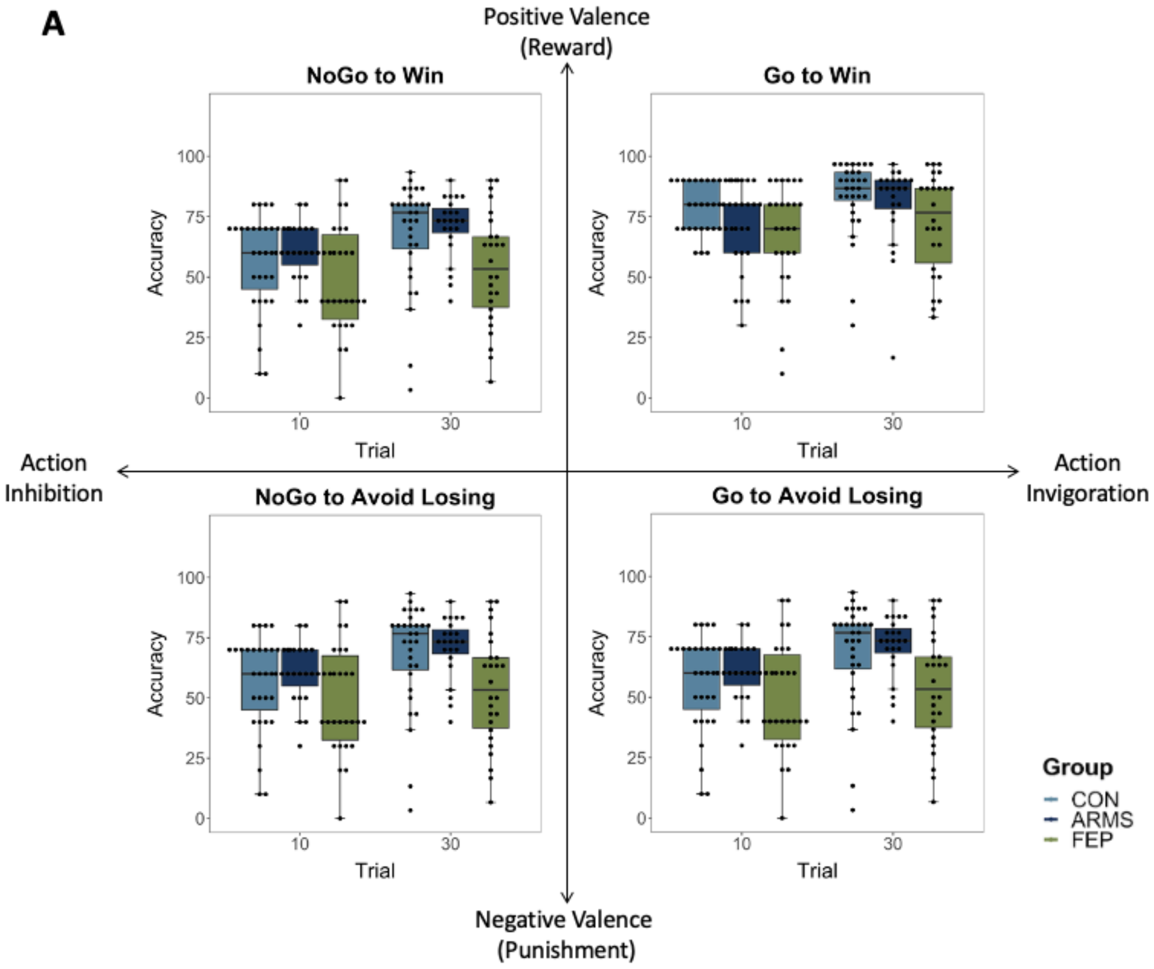


*Note.* A. shows individual learning rates at trial 10 and 30 by group and trial type.

2. **Logistic Regression for ROC analysis based on AI parameters**

***CONvsARMS***

```
Call:
glm(formula = Group_bin ~ PrvW + PrvGgW + PrvNGgAL + b_r + frg +
    precAlpha + F, family = binomial(), data = CONvsARMS, na.action = na.exclude)

Deviance Residuals:
    Min       1Q   Median       3Q      Max
-1.4028  -1.0289  -0.6893   1.1465   1.8227

Coefficients:
             Estimate Std. Error z value Pr(>|z|)
(Intercept)  32.41166   37.11644   0.873    0.383
PrvW         -3.75166    7.04545  -0.532    0.594
PrvGgW        3.36181    5.62615   0.598    0.550
PrvNGgAL    -12.89157    8.44087  -1.527    0.127
b_r          -5.58768    7.13245  -0.783    0.433
frg           0.96671    2.83427   0.341    0.733
precAlpha     0.21496    1.02811   0.209    0.834
F            -0.02125    0.03026  -0.702    0.482

(Dispersion parameter for binomial family taken to be 1)

    Null deviance: 73.670  on 53  degrees of freedom
Residual deviance: 66.979  on 46  degrees of freedom
AIC: 82.979

Number of Fisher Scoring iterations: 5
```

***CONvsFEP***

```
Call:
glm(formula = Group_bin ~ PrvW + PrvGgW + PrvNGgAL + b_r + frg +
    precAlpha + F, family = binomial(), data = CONvsFEP_complete)

Deviance Residuals:
    Min       1Q   Median       3Q      Max
-1.6441  -0.8247  -0.4270   0.9686   2.3641

Coefficients:
            Estimate Std. Error z value Pr(>|z|)
(Intercept) 22.52886   35.78976   0.629   0.5290
PrvW         8.10339    8.82458   0.918   0.3585
PrvGgW       1.39980    4.62518   0.303   0.7622
PrvNGgAL     7.31184    8.09227   0.904   0.3662
b_r         -7.01987    6.92313  -1.014   0.3106
frg          4.20751    2.35934   1.783   0.0745 .
precAlpha    0.12014    1.07748   0.112   0.9112
F           -0.02400    0.03032  -0.792   0.4286
---
Signif. codes:  0 ‘***’ 0.001 ‘**’ 0.01 ‘*’ 0.05 ‘.’ 0.1 ‘ ’ 1

(Dispersion parameter for binomial family taken to be 1)

    Null deviance: 75.353  on 54  degrees of freedom
Residual deviance: 59.439  on 47  degrees of freedom
AIC: 75.439

Number of Fisher Scoring iterations: 5
```

***ARMSvsFEP***

```
Call:
glm(formula = Group_bin ~ PrvW + PrvGgW + PrvNGgAL + b_r + frg + 
    precAlpha + F, family = binomial(), data = ARMSvsFEP_complete)

Deviance Residuals: 
    Min       1Q   Median       3Q      Max  
-2.1154  -0.9505   0.1073   0.8742   1.8633  

Coefficients:
            Estimate Std. Error z value Pr(>|z|)  
(Intercept) 23.02057   76.42254   0.301   0.7632  
PrvW        16.35588    9.81616   1.666   0.0957 .
PrvGgW       3.55991    6.49684   0.548   0.5837  
PrvNGgAL    27.15656   13.12896   2.068   0.0386 *
b_r         -8.83041   15.14641  -0.583   0.5599  
frg          7.72238    3.34782   2.307   0.0211 *
precAlpha   -1.62976    1.23701  -1.318   0.1877  
F            0.05451    0.03699   1.474   0.1405  
---
Signif. codes:  0 '***' 0.001 '**' 0.01 '*' 0.05 '.' 0.1 ' ' 1

(Dispersion parameter for binomial family taken to be 1)

    Null deviance: 65.135  on 46  degrees of freedom
Residual deviance: 47.515  on 39  degrees of freedom
AIC: 63.515

Number of Fisher Scoring iterations: 6
```

### 3. Logistic Regression for ROC analysis based on performance

***CONvsARMS***

```
Call:
glm(formula = Group_bin ~ cue1 + cue2 + cue3 + cue4, family = binomial(), 
    data = CONvsARMS, na.action = na.exclude)

Deviance Residuals: 
   Min      1Q  Median      3Q     Max  
-1.436  -1.057  -0.925   1.263   1.408  

Coefficients:
              Estimate Std. Error z value Pr(>|z|)
(Intercept)  0.5810283  1.8714394   0.310    0.756
cue1        -0.0087664  0.0184216  -0.476    0.634
cue2         0.0140753  0.0195842   0.719    0.472
cue3         0.0001295  0.0117396   0.011    0.991
cue4        -0.0148708  0.0193985  -0.767    0.443

(Dispersion parameter for binomial family taken to be 1)

    Null deviance: 73.670  on 53  degrees of freedom
Residual deviance: 72.191  on 49  degrees of freedom
AIC: 82.191

Number of Fisher Scoring iterations: 4
```

#### *CONvsFEP*

```
Call:
glm(formula = Group_bin ~ cue1 + cue2 + cue3 + cue4, family = binomial(),
    data = CONvsFEP_complete)

Deviance Residuals:
    Min       1Q   Median       3Q      Max
-1.5835  -0.7491  -0.4871   0.8127   2.1886

Coefficients:
            Estimate Std. Error z value Pr(>|z|)
(Intercept)  4.99015    1.72804   2.888  0.00388 **
cue1         0.01586    0.02303   0.689  0.49114
cue2        -0.05236    0.02291  -2.286  0.02228 *
cue3         0.03312    0.02053   1.613  0.10673
cue4        -0.07187    0.02586  -2.779  0.00546 **
---
Signif. codes:  0 ‘***’ 0.001 ‘**’ 0.01 ‘*’ 0.05 ‘.’ 0.1 ‘ ’ 1

(Dispersion parameter for binomial family taken to be 1)

    Null deviance: 75.353  on 54  degrees of freedom
Residual deviance: 57.739  on 50  degrees of freedom
AIC: 67.739

Number of Fisher Scoring iterations: 4
```

#### *ARMSvsFEP*

```
Call:
glm(formula = Group_bin ~ cue1 + cue2 + cue3 + cue4, family = binomial(),
    data = ARMSvsFEP_complete)

Deviance Residuals:
    Min       1Q   Median       3Q      Max
-1.7968  -0.7651   0.3848   0.8069   2.0839

Coefficients:
             Estimate Std. Error z value Pr(>|z|)
(Intercept)  4.228671   1.798851   2.351   0.0187 *
cue1         0.019710   0.023932   0.824   0.4102
cue2        -0.057945   0.025151  -2.304   0.0212 *
cue3         0.003583   0.016818   0.213   0.8313
cue4        -0.032162   0.018409  -1.747   0.0806 .
---
Signif. codes:  0 ‘***’ 0.001 ‘**’ 0.01 ‘*’ 0.05 ‘.’ 0.1 ‘ ’ 1

(Dispersion parameter for binomial family taken to be 1)

    Null deviance: 65.135  on 46  degrees of freedom
Residual deviance: 51.516  on 42  degrees of freedom
AIC: 61.516

Number of Fisher Scoring iterations: 4
```

4. **Logistic Regression for ROC analysis based on AI parameters and performance**

***CONvsARMS***

```
Call:
glm(formula = Group_bin ~ PrvW + PrvGgW + PrvNGgAL + b_r + frg +
    precAlpha + F + cue1 + cue2 + cue3 + cue4, family = binomial(),
    data = CONvsARMS, na.action = na.exclude)

Deviance Residuals:
   Min      1Q  Median      3Q     Max
-1.615  -1.018  -0.429   1.038   1.916

Coefficients:
             Estimate Std. Error z value Pr(>|z|)
(Intercept)  18.21332   42.28023   0.431    0.667
PrvW          9.49428   10.61228   0.895    0.371
PrvGgW       16.33840   10.58604   1.543    0.123
PrvNGgAL    -11.10886    9.20477  -1.207    0.227
b_r          -8.18965    8.18003  -1.001    0.317
frg           4.69681    3.63867   1.291    0.197
precAlpha    -0.30825    1.11603  -0.276    0.782
F            -0.06341    0.04077  -1.555    0.120
cue1          0.01077    0.02414   0.446    0.655
cue2          0.02397    0.02722   0.881    0.379
cue3          0.02838    0.02243   1.265    0.206
cue4          0.05957    0.04248   1.402    0.161

(Dispersion parameter for binomial family taken to be 1)

    Null deviance: 73.670  on 53  degrees of freedom
Residual deviance: 63.166  on 42  degrees of freedom
AIC: 87.166

Number of Fisher Scoring iterations: 5
```

***CONvsFEP***

```
Call:
glm(formula = Group_bin ~ PrvW + PrvGgW + PrvNGgAL + b_r + frg +
    precAlpha + F + cue1 + cue2 + cue3 + cue4, family = binomial(),
    data = CONvsFEP_complete)

Deviance Residuals:
    Min       1Q   Median       3Q      Max
-2.0505  -0.6811  -0.1304   0.6380   2.3224

Coefficients:
             Estimate Std. Error z value Pr(>|z|)
(Intercept)  7.954347  33.885905   0.235  0.81441
PrvW        -3.649653  11.528860  -0.317  0.75157
PrvGgW      -7.568022  11.494608  -0.658  0.51028
PrvNGgAL    38.491045  17.002737   2.264  0.02359 *
b_r         -3.994159   6.047426  -0.660  0.50895
frg         -0.814419   4.054929  -0.201  0.84082
precAlpha    2.691696   1.586058   1.697  0.08968 .
F           -0.051039   0.040076  -1.274  0.20282
cue1         0.002738   0.037951   0.072  0.94248
cue2        -0.024544   0.031694  -0.774  0.43869
cue3         0.041373   0.032857   1.259  0.20798
cue4        -0.151578   0.056887  -2.665  0.00771 **
---
Signif. codes:  0 ‘***’ 0.001 ‘**’ 0.01 ‘*’ 0.05 ‘.’ 0.1 ‘ ’ 1

(Dispersion parameter for binomial family taken to be 1)

    Null deviance: 75.353  on 54  degrees of freedom
Residual deviance: 46.694  on 43  degrees of freedom
AIC: 70.694

Number of Fisher Scoring iterations: 6
```

### *ARMSvsFEP*

```
Call:
glm(formula = Group_bin ~ PrvW + PrvGgW + PrvNGgAL + b_r + frg +
    precAlpha + F + cue1 + cue2 + cue3 + cue4, family = binomial(),
    data = ARMSvsFEP_complete)

Deviance Residuals:
     Min        1Q    Median        3Q       Max
-2.17560  -0.57094   0.01775   0.63693   1.54798

Coefficients:
            Estimate Std. Error z value Pr(>|z|)
(Intercept) 18.98202   91.84037   0.207   0.8363
PrvW         5.78166   13.25534   0.436   0.6627
PrvGgW       5.95269   14.47379   0.411   0.6809
PrvNGgAL    30.76723   18.43868   1.669   0.0952 .
b_r         -4.26327   18.22355  -0.234   0.8150
frg          8.18944    5.52434   1.482   0.1382
precAlpha   -3.24423    1.95411  -1.660   0.0969 .
F            0.15799    0.07906   1.998   0.0457 *
cue1         0.06716    0.04683   1.434   0.1515
cue2        -0.13014    0.06798  -1.915   0.0555 .
cue3         0.04154    0.04406   0.943   0.3458
cue4        -0.08345    0.05325  -1.567   0.1171
---
Signif. codes:  0 ‘***’ 0.001 ‘**’ 0.01 ‘*’ 0.05 ‘.’ 0.1 ‘ ’ 1

(Dispersion parameter for binomial family taken to be 1)

    Null deviance: 65.135  on 46  degrees of freedom
Residual deviance: 37.610  on 35  degrees of freedom
AIC: 61.61

Number of Fisher Scoring iterations: 6
```

### 5. Correlation of model parameter, model fit and clinical scores by group

**Supplementary Figure 2:** *Heatmaps AI Parameters and Clinical Parameters*

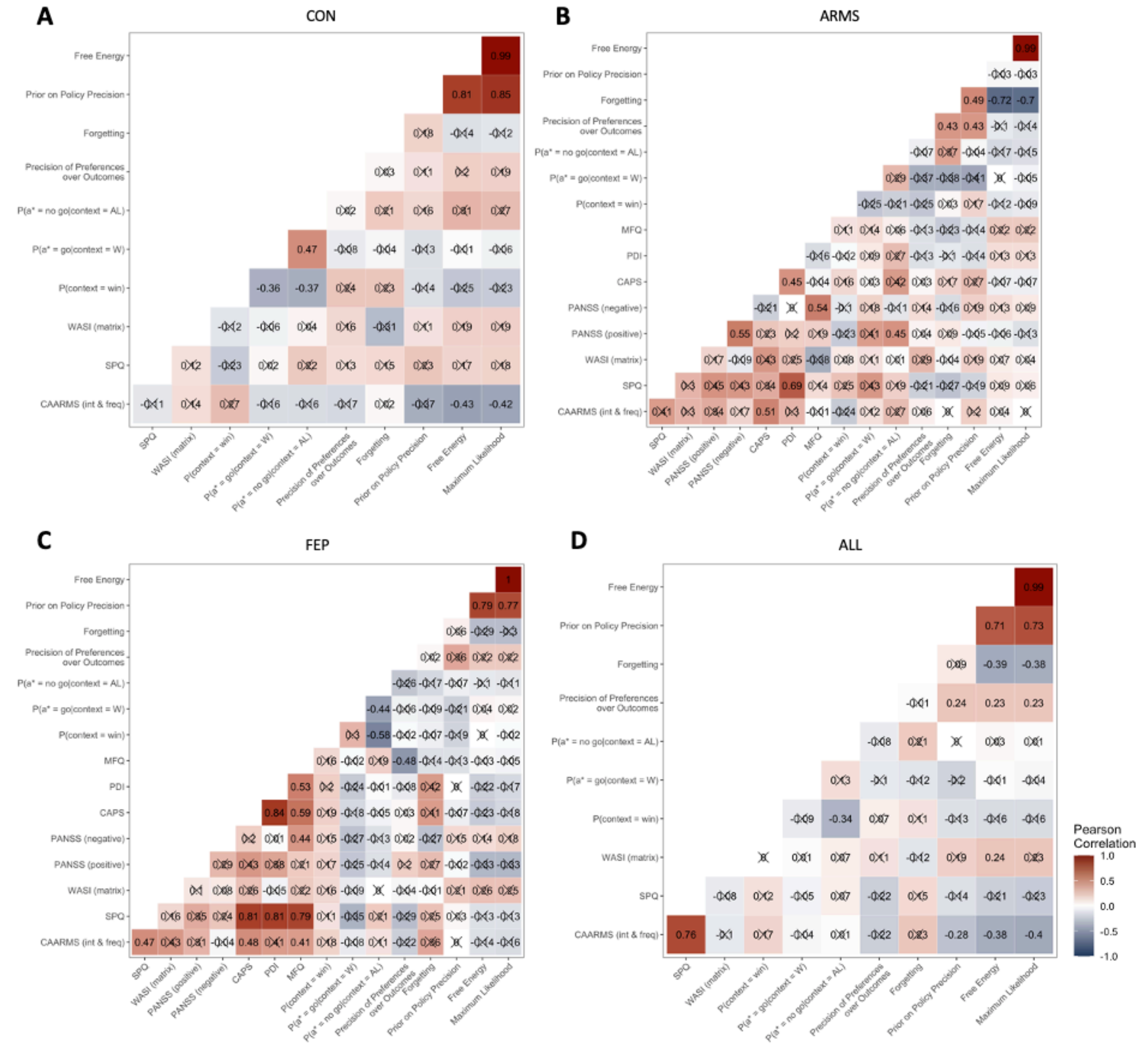


*Note. Correlations of modelling parameters and clinical scores across Controls (A), ARMS (B), FEP (C) and across all participants (D).*

### Model fit – comparing learners and non-learners in the final 20 trials

To investigate whether the AI model provides a better fit for the better-performing individuals, we used  the fraction of correct responses in the final 20 trials, as previous research observed that performance approximately stabilizes by this time point (Adams et al., 2020; Moutoussis et al., 2019).

**Comparison of Model Fit Parameters**

**Supplementary Figure 3:** *Comparison of Model Fit Parameters*

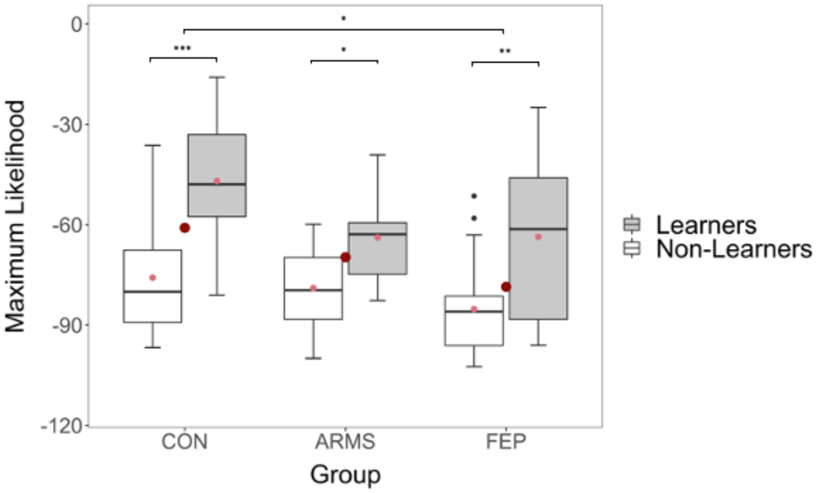


*Note. ANOVA analysis of group differences of maximum likelihood measures for learners compared to non-learners. Significant results from the Tukey corrected post hoc analyses are shown (*p < 0.05, **p < 0.01, ***p< 0.001). CON (Healthy Controls); ARMS (At-Risk for Mental Health); FEP (First-Episode Psychosis). Box plots show the median as horizontal bars, mean as small red dot, group mean as large red dot, interquartile range as whiskers.*

Based on the findings of Adams et al. (2020), we compared the model fit as assessed by maximum likelihood measures for learners vs. non-learners within and between groups. A two-way ANOVA showed significant main effects for both variables group ($F(2,74) = 4.21$, $p = .19$) and learners ($F(1,74) = 29.33$, $p < .001$) without a significant interaction effect. Post-hoc Tukey tests revealed that maximum likelihood values were significant higher for healthy controls compared to FEP individuals (2.71, 95% CI [1.53, 24.5], $p = .023$) and that the model fit was in general better for learners compared to non-learners (21.9, 95% CI [13.8, 29.9], $p < .001$).
